# Implementation Adherence and Operational Challenges of Rectal Artesunate for Severe Malaria in Zambia: A Mixed-Methods Study

**DOI:** 10.64898/2026.05.20.26353691

**Authors:** Andrew Andrada, Ernest Chanda, Isabelle Smith, Olivia Sam, Irene Kyomuhangi, John Miller, Kafula Silumbe, Cathy Green, Hans Rietveld, Stephen Bwalya, Busiku Hamainza, Japhet Chiwaula, Jayne Webster, Yazoume Ye, Eva Silvestre, Ruth A Ashton, Thomas P. Eisele

**Affiliations:** Center for Applied Malaria Research and Evaluation, Tulane University School of Public Health and Tropical Medicine, New Orleans, LA, United States; PATH-Malaria Control and Elimination Partnership in Africa (MACEPA), Chainama Hospital College Grounds, Lusaka, Zambia; Tulane University School of Public Health and Tropical Medicine, New Orleans, LA, United States; Bennett Institute for Applied Data Science, Oxford University, Oxford, UK; Transaid, 137 Euston Road, London NW1 2AA, England; Medicines for Malaria Venture, Meyrin, Switzerland; National Malaria Elimination Centre, Zambia Ministry of Health, Lusaka, Zambia; Disease Control Department, London School of Hygiene and Tropical Medicine, London, UK; CESMEL Health, Bowie, MD, United States

## Abstract

Rectal artesunate (RAS) is a pre-referral intervention recommended for children with suspected severe malaria in remote settings where injectable treatment is not readily available. Although clinical trials have demonstrated efficacy, less is known about the behavioural and health system factors influencing effectiveness under routine conditions. A convergent parallel mixed-methods design was used to assess implementation of Zambia’s RAS intervention package across three districts: Serenje, Chama, and Mwinilunga. A retrospective case-tracking investigation of all 300 children with suspected severe malaria recorded by community health workers (CHWs) assigned to study facilities examined progression and attrition across the severe malaria care cascade. In-depth interviews and focus group discussions with caregivers, CHWs, and other stakeholders explored barriers and facilitators influencing progression. Among 300 enrolled children, early attrition occurred due to negative rapid diagnostic test results. Of 239 RDT-positive children, 218 (91.2%) received RAS. Referral completion was lower; among 261 children referred and followed up at health facilities, 209 (80.1%) were confirmed to have completed referral. Of 186 children diagnosed with severe malaria at the facility, 167 (89.8%) received both injectable artesunate and follow-on artemether–lumefantrine. Patterns of disengagement varied by district, with Serenje demonstrating the most consistent progression, Chama experiencing the largest drop-off at RAS administration, and Mwinilunga showing the lowest completion of follow-on treatment. Qualitative findings revealed strong community appreciation for RAS despite stockouts, alongside social and behavioural barriers, including gendered responsibilities, transport challenges, and confusion following symptom improvement, that discouraged referral completion. RAS can be a life-saving intervention when embedded within strong health systems and community structures. Zambia’s experience underscores the need for comprehensive implementation strategies that extend beyond drug distribution to include sustained CHW training, reliable commodity supply, functional referral systems, and meaningful caregiver engagement.

## Background

In 2024, an estimated 610,000 malaria deaths occurred globally, the vast majority among young children in sub-Saharan Africa [1]. Children are among the most vulnerable and their risk of malaria mortality is high due to their lack of acquired immunity [2]. Once uncomplicated malaria progresses to severe disease, it can rapidly become fatal within hours if appropriate treatment is not obtained [2]. Ensuring that a child with severe malaria living in a rural, and sometimes hard-to-reach, area receives timely, appropriate care remains a significant operational and behavioural challenge. Access to first-line treatment with injectable antimalarials is often limited, with health facilities taking hours or even days to reach by foot. To bridge this gap, rectal artesunate (RAS) capsules were introduced as a life-saving pre-referral intervention to stabilize severely ill children and buy them time to reach critical care. Randomized trials have demonstrated that RAS significantly reduces severe complications among children who reached a health facility more than six hours after administration compared to those who received no pre-referral RAS [3]. Based on these findings, the World Health Organization (WHO) recommends RAS in settings where injectable antimalarials, part of the treatment for severe malaria, are unavailable close to communities, and its implementation has slowly expanded across sub-Saharan Africa [4, 5].

However, while these studies established the clinical efficacy of RAS, they did not evaluate the behavioural factors and health system supports required to complete the full treatment pathway for severe malaria. The cascade of care for severe malaria consists of four critical steps: 1) treatment-seeking for the life-threatening condition, 2) diagnosis and administration of RAS at the community-level, 3) timely completion of referral at a health facility, and 4) complete treatment with injectable artesunate (Inj AS) for at least 24 hours plus a full 3-day course of an oral artemisinin-based combination therapy (ACT), such as artemether lumefantrine (AL), as soon as the child can swallow and retain the medication (Fig 1). Each step is necessary to reduce the risk of permanent disability and/or death. The journey for children in remote settings to complete the cascade of care relies on a combination of social, behavioural, and health system factors. These include caregiver decision-making and awareness, financial and geographic access to referral facilities, trust in the health providers, availability of essential commodities (such as RAS, AL, and injectable AS), and competence and responsiveness of community health workers (CHWs) and health facility staff [6–9]. To improve RAS effectiveness these factors should be accounted for in each country’s implementation plan.

**Fig 1.**
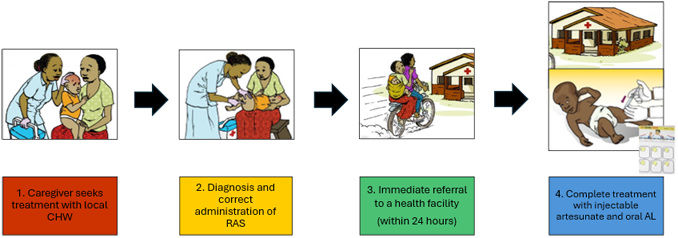
Cascade of care for severe malaria. Note: CHW = community health worker, RAS = rectal artesunate, AL = artemether-lumefantrine. Images were adapted from the Medicines for Malaria Venture *Rectal Artesunate User Guide* (2019) [22]

Despite its increasing use, research on RAS implementation has mainly been quantitative, leaving critical gaps in the evidence base on how real-world operational, social, and behavioural factors influence completion of the cascade of care for severe malaria. One of the largest studies evaluating real-world RAS implementation, the Community Access to Rectal Artesunate for Malaria (CARAMAL) project, was conducted in the Democratic Republic of Congo, Nigeria, and Uganda [10]. This large-scale observational study identified major health system and implementation barriers. Notably, CARAMAL results showed that <10% of children who received stand-alone RAS went on to receive appropriate treatment at referral facilities [11]. The authors hypothesized that caregivers, seeing their child’s condition improve after receiving RAS, no longer perceived referral as necessary—leading to incomplete referral and unexpectedly high mortality among RAS recipients [11, 12].

Critics of the CARAMAL project’s results have argued that several biases inherent to its observational design were not adequately addressed, raising concerns about the interpretation of its findings [13–15]. Nevertheless, these results highlight the urgent need to understand and address the health system, social, behavioural, and systemic factors influencing each step of the RAS cascade. Qualitative research to date has primarily explored the community acceptability and understanding of RAS [16–18]. A gap remains in identifying the challenges that key actors along the cascade of care encounter when treating a child with severe malaria.

Zambia provides a valuable case study for RAS implementation, as the intervention has been progressively scaled up since 2017 with the goal of reducing rural malaria mortality [19]. The Zambia National Malaria Elimination Centre (NMEC) and its partners initially piloted RAS in Serenje District and later expanded it to ten districts across the country [20, 21]. Zambia’s RAS intervention package comprised more than just the drug itself—it included the provision of RAS commodities, comprehensive training for CHWs and facility staff on severe malaria case management, supportive supervision, and a referral system strengthened through emergency transport schemes and community-based support mechanisms such as savings groups and food banks to assist families with referral costs. This phased rollout provided a unique opportunity to evaluate implementation strengths, challenges, and contextual differences across multiple settings. While previous research on RAS in Zambia has demonstrated its potential effectiveness, these evaluations were conducted under ideal conditions with continuous external support and funding to ensure RAS was implemented as planned [20, 21]. This study is complementary as it examines how the RAS intervention package is functioning under routine conditions several years post-pilot, without the same level of external support.

The study assessed Zambia’s current RAS intervention package implementation by examining how well children with severe malaria progressed through the cascade of care, alongside evaluating the service availability and readiness of CHWs, health facilities, and emergency transport systems. By also exploring the perspectives of caregivers and health system actors, this research aimed to identify health system and social, behavioural, and practical barriers that influence treatment completion. These insights are critical for informing program strategies to maximize the life-saving potential of RAS and avoid unintended risks from incomplete care.

## Methods

### Study design and study sites

This study used a convergent parallel mixed-methods design, in which quantitative tracking of severe malaria cases and qualitative interviews were conducted during the same period and integrated during analysis to generate a comprehensive understanding of implementation adherence and contextual determinants. The study examined compliance to three of the four critical steps in the severe malaria care cascade across three Zambian districts with different levels of RAS implementation: Serenje (pilot site, launched December 2018), Chama (first scale-up phase, July 2019), and Mwinilunga (second scale-up phase, November 2020).

Two referral health facilities per district (six in total) were selected based on severe malaria case burden and recommendations from the local health team regarding RAS implementation. These facilities were either rural health posts or rural health centres. The first step—treatment seeking for severe malaria at the community level—could not be quantified as the study design only captured children whose caregivers sought care from a CHW.

During the pilot phase in Serenje, the RAS package was introduced with intensive support, including comprehensive CHW and facility staff training, reliable RAS supply, functional emergency transport systems (ETS; e.g., bicycle ambulances), and strong community support systems such as savings schemes and food banks to assist families with referral costs. CHWs were also trained to address gender-based barriers (e.g. gender-based violence; lack of male involvement in children’s health) that affected timely treatment seeking and to identify and include socially excluded individuals in their activities. Supervision was frequent, and facility readiness to provide injectable artesunate and ACTs was reinforced [21]. As implementation expanded to Chama in the first scale-up phase, the level of support was intentionally reduced to approximate routine health system conditions [20]. Trainings were shorter, refresher trainings less frequent, and external supervision lighter. While CHWs were trained and RAS was available, community support systems were less extensive and more inconsistently maintained. In Mwinilunga, added during the second scale-up phase, support was further reduced. Most CHWs had received some training in RAS, but refresher trainings were rare, stock-outs more common, no functional emergency transport system was in place, and community support structures such as savings schemes or food banks were largely absent.

## Data collection

### Adapted service availability and readiness assessments

Adapted Service Availability and Readiness Assessments (aSARAs), based on the WHO’s Service Availability and Readiness Assessment (SARA) tool, were tailored to the RAS intervention package and collected data on CHWs, health facility staff, and emergency transport providers [23]. These aSARAs were conducted to understand CHW capacity to provide RAS, determine emergency referral transportation availability, and evaluate referral facility readiness to provide complete management of severe malaria [23].

### Severe malaria case investigation

All suspected severe malaria cases seen by CHWs assigned to the study health facilities between 01/01/2022 and 31/05/2023 were retrospectively included in this study. Each case was tracked to assess progression through the cascade of care, with data collected at both the community and facility-levels. CHW logbooks and facility records were first accessed by the study team beginning 26/06/2023. During data abstraction, study staff had access to identifiable information contained in CHW logbooks and facility registers (including child name and village of residence) for the purpose of record linkage and follow-up. Identifiable information was used solely to verify referral completion and conduct household follow-up visits and was removed prior to analysis. All analytic datasets were de-identified, and no identifying information was retained in the final dataset used for statistical analysis.

The primary objective was to determine where, why, and how children with suspected severe malaria either completed or failed to complete referrals, and to quantify the proportion of cases achieving each of the three measurable steps in the cascade of care. Data were obtained from CHW logbooks, health facility records, and household follow-up visits with the families of identified children

CHW logbooks provided patient demographic and clinical information including age, sex, residence, date of visit, symptoms, diagnosis, treatment, and referral details. Once a roster of patients was compiled electronically, the study team conducted follow-up at two levels. Facility follow-up involved visits to referral facilities to verify referral completion through review of facility registers and health worker confirmation. Data obtained included diagnosis and treatment provided. Community follow-up consisted of home visits to verify the child’s survival status, confirm referral completion if not recorded in facility data, and document transport used for referral and receipt of injectable AS followed by a full course of oral AL.

Not all children could be traced during follow-up due to migration or remote locations; therefore, survival outcomes and treatment completion were calculated only among children with verified follow-up. All data were collected using CommCare (Dimagi, Cambridge MA), a digital data collection application installed on Android mobile devices.

### In-depth interviews and focus group discussions

In-depth interviews (IDIs) were conducted with key stakeholders across the cascade of care, including caregivers, CHWs, health facility staff, district, and provincial health teams, and NMEC staff. Recruiting for qualitative interviews began on 12/07/2023. Caregivers were sequentially sampled for IDIs based on referral completion within the selected districts, while other key stakeholders were identified with assistance from District Health Management Teams (DHMTs). Focus group discussions were held with community members from each selected health facility catchment area, with participants identified in collaboration with the DHMTs. No incentives were provided for respondents throughout the qualitative data collection.

All qualitative data collectors were trained in qualitative data collection procedures, interview guides, and ethics. Interview guides were developed to reflect the assumed steps and mechanisms of the severe malaria cascade of care and study outcomes. Ethical approval was obtained from the University of Zambia, Tulane University, and PATH IRBs, as detailed in the Ethics section. These guides were first developed in English and were reviewed by the study team prior to pretesting. Revised guides were then shared with key stakeholders prior to the data collection to obtain their feedback. Once finalized, the interview guides were translated into the local dialects of Bemba, Lunda, and Nsenga, for the selected districts, Serenje, Mwinilunga, and Chama, respectively. These translations were then reviewed and back translated by separate translators for verification prior to data collection.

IDIs explored experiences and perceptions during the critical steps in the cascade of care for severe malaria, which include correct diagnosis and administration of RAS, immediate referral, and prompt and complete treatment of referred children with injectable artesunate followed by oral AL. All participants were aged 16 years (the age of consent in Zambia) or older. Written informed consent was obtained and participants were asked again prior to data collection for their verbal consent, which was documented by the interviewer on a study consent form approved by the institutional review boards. Digital recorders were used to record each interview. Zoom was also used when respondents were unable to meet in person. Interviewers kept detailed notes and completed interview summary forms that were shared with the study team.

### Data management and analysis

#### Severe malaria case investigation

Data were collected on children with suspected severe malaria that were seen by a CHW. Each case was investigated to generate proportions of severe malaria cases achieving each of the three critical steps in the cascade of care. These proportions were calculated at the health facility level and then aggregated to provide district-level estimates, allowing comparison of performance across sites. Each critical step in the cascade of care was assessed for variation in loss at each step across the different districts/facilities, assessing where children with severe malaria were most likely to drop out of the care cascade.

#### Qualitative interview analysis

The audio recordings from IDIs and FGDs were simultaneously transcribed and translated into English by an independent local transcription service in Lusaka. After transcription, the research team reviewed the transcripts and imported them into NVivo 15 for coding and analysis [24]. To maintain confidentiality, each transcript was assigned a unique anonymized identifier indicating respondent type and district.

A thematic analysis approach was applied, using the severe malaria cascade of care as the primary analytic framework. The research team first familiarized themselves with the transcripts with an initial read-through. An initial coding framework was developed with the cascade’s key steps as major coding nodes. Each transcript was coded using the predefined framework and respondent type. Inductive codes were added as child codes within the relevant cascade step when new concepts emerged. Two US-based research assistants primarily coded data, and three coding validation sessions were held with the study team to ensure consistency of code interpretation. As coding progressed across respondent types and districts, the study team assessed data saturation to monitor whether new themes continued to emerge. After coding was completed, the frequency of each code and its associated text segments was examined to assess salience of facilitator and barrier themes across districts and respondent types. This helped identify which challenges were most commonly experienced and where patterns in the cascade of care consistently broke down.

Comprehensive member checking with the interview participants was not feasible. Instead, the local study manager, who oversaw the data collection and facilitated several interviews, validated the emergent themes. This involved presenting a concise summary of the key findings, discussing each thematic category, and addressing potential gaps or misinterpretations. The study manager’s insights informed final revisions, ensuring that the analysis accurately reflected the local context and participant perspectives. Finally, the qualitative findings were used to refine the assumed steps and mechanisms underlying the severe malaria cascade of care, incorporating additional factors influencing RAS implementation.

## Results

### Enrolment, follow-up, and cascade attrition

A total of 300 children with suspected severe malaria were identified in CHW logbooks across the three study districts: 118 in Serenje, 117 in Chama, and 65 in Mwinilunga (Fig 2). Follow-up was conducted at both health facilities and in communities to verify referral completion, treatment received, and survival status. Health facility follow-up was attempted for 261 children (87.0%), of whom 209 (80.1%) were found in facility records. Community follow-up was completed for 202 children (67.3%), with families located in 83.2% of attempted visits.

**Fig 2.**
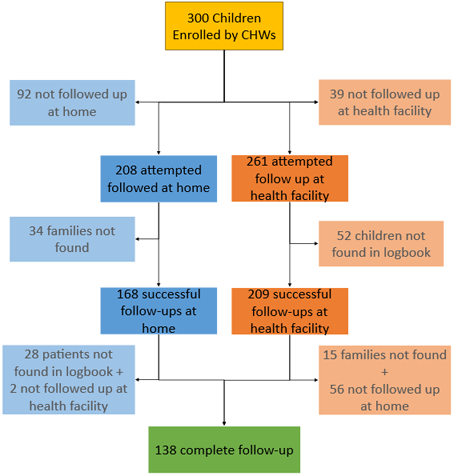
Case follow-up flow diagram for enrolled children with suspected severe malaria across the study districts

Because follow-up was not possible for all children—primarily due to late enrollment as teams moved between sites and the remoteness of some households—full outcome information was not available for every case. Children with missing follow-up data were retained in the cascade for all steps where verified information existed, and additional details on follow-up completeness and loss to follow-up by district are provided in S1 Table.

Across the severe malaria cascade of care, substantial attrition occurred at multiple steps following initial CHW enrolment (Fig 3). While most children received an RDT (95.3%), only 72.7% were administered RAS, and fewer than 70% completed referral to a health facility. Attrition continued at the facility level, with 61.3% receiving injectable artesunate and only 56.3% completing treatment with artemether–lumefantrine, highlighting progressive losses across the continuum of care.

**Fig 3.**
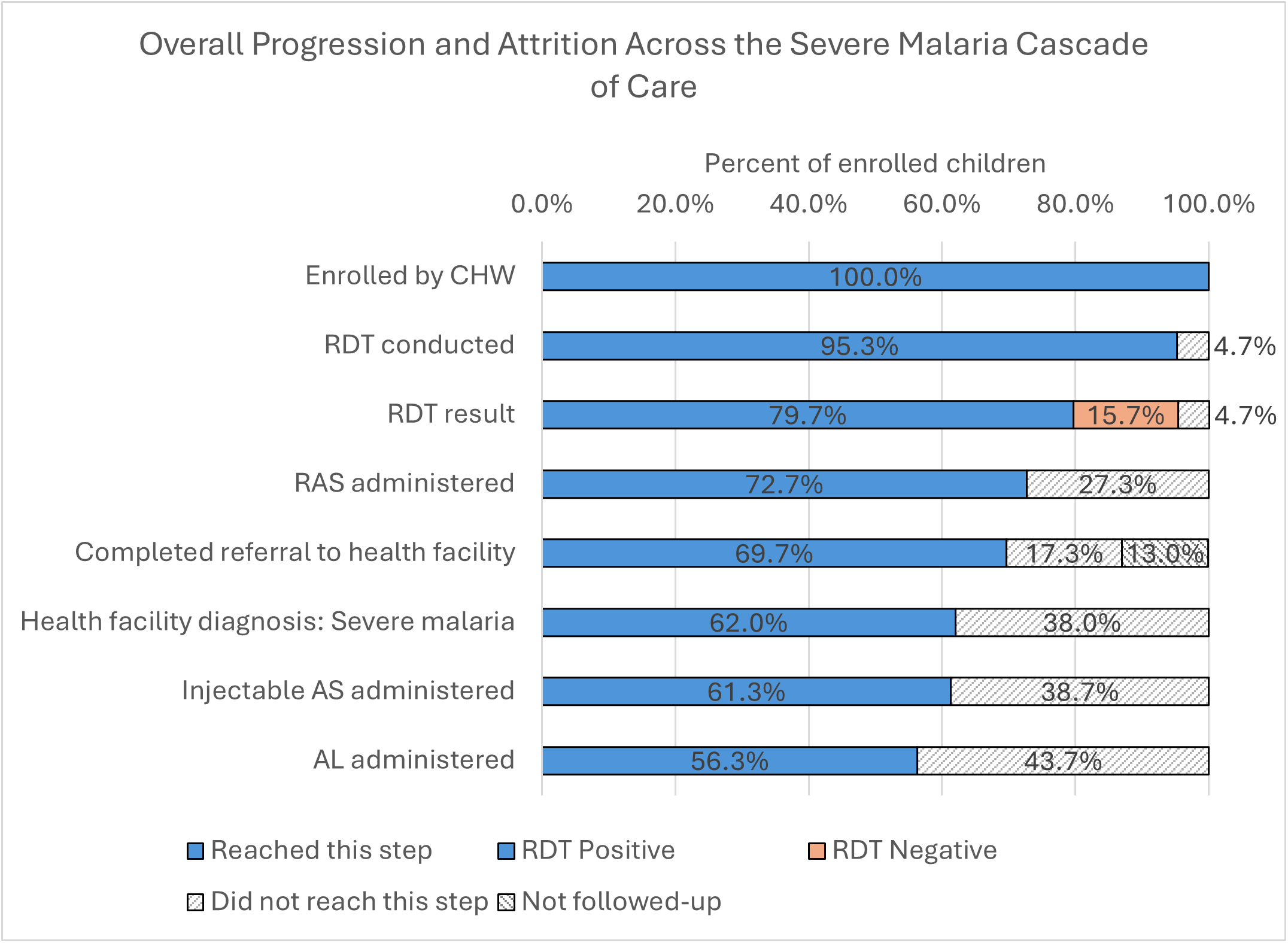
Overall progression and attrition across the severe malaria cascade of care. At each step, “Did not reach this step” represents children who either were not eligible to progress to that step *or* whose progression could not be assessed due to lack of followup

### Study patient characteristics by RAS administration

Of the 300 children enrolled with suspected severe malaria, 218 (72.7%) were administered RAS (Table 1). There were no statistically significant differences in gender and age between those that received RAS and those that did not. However, children who refused to eat/drink (p<0.001), vomited (p=0.012), and/or had difficulty waking up (p=0.007) during the CHW visit were more likely to receive RAS than those presenting with other symptoms. RAS was also more likely to be administered during the rainy season (November to April) (p=0.037). There were also significant differences across the districts (p<0.001), with nearly half the children receiving RAS residing in Serenje (47.7%) followed by Chama (29.8%) and Mwinilunga (22.5%). In contrast, among children that did not receive RAS, most were from Chama (63.4%), followed by Mwinilunga (19.5%), then Serenje (17.1%).

**Table 1.** Study patient characteristics by RAS administration.

| Background Characteristic | RAS<br>N = 218<sup>1</sup> | No RAS<br>N = 82<sup>1</sup> | p-value<sup>2</sup> |
| --- | --- | --- | --- |
| **Gender** |  |  | 0.9 |
| Female | 110 (50.5%) | 40 (48.8%) |  |
| Male | 108 (49.5%) | 42 (51.2%) |  |
| **Mean age in years (SD)** | 2 (2) | 2 (2) | >0.9 |
| **Observed symptoms by CHW** |  |  |  |
| Fever | 208 (95.4%) | 80 (97.6%) | 0.5 |
| Fitting | 129 (59.2%) | 52 (63.4%) | 0.6 |
| Refuse to eat/drink | 100 (45.9%) | 8 (9.8%) | **<0.001** |
| Vomiting | 97 (44.5%) | 23 (28.0%) | **0.012** |
| Difficulty in waking up | 23 (10.6%) | 1 (1.2%) | **0.007** |
| **Enrolment location** |  |  | 0.8 |
| With CHW | 85 (39.0%) | 32 (39.0%) |  |
| From CHW logbook | 117 (53.7%) | 42 (51.2%) |  |
| Referral slip | 16 (7.3%) | 8 (9.8%) |  |
| **District** |  |  | **<0.001** |
| Chama | 65 (29.8%) | 52 (63.4%) |  |
| Mwinilunga | 49 (22.5%) | 16 (19.5%) |  |
| Serenje | 104 (47.7%) | 14 (17.1%) |  |
| **Rainy season\*** | 129 (59.2%) | 37 (45.1%) | **0.037** |
| <sup>1</sup>n (%); Mean (SD) |  |  |  |
| <sup>2</sup>Fisher's exact test (categorical variables); Welch Two Sample t-test (age) |  |  |  |
| *November-April |  |  |  |

### Cascade of care for severe malaria

#### Step 1: Caregiver treatment-seeking (Qualitative only)

Caregivers must first decide whether, when and where to seek care. Many chose CHWs for their accessibility and trusted skills: *“Even when we go at night, they wake up and attend to our children”. (FGD Participant 1, Serenje)*. However, some caregivers bypassed CHWs and went directly to facilities, often due to past experiences with CHW stockouts or doubts about their ability to manage severe cases. Delays were also common when symptoms appeared mild or at night: *“You find that the child is sick in the evenings and they will wait until morning… This delay puts the life of the child at risk”. (Caregiver 1 IDI, Chama)*. In some cases, distrust in diagnostic results led families to use traditional medicine instead: *“Last time I went there it was negative… instead I will just do the traditional medicine”. (FGD Participant 2, Mwinilunga)*.

Although no quantitative data were collected for this step, facility records occasionally showed severe malaria cases without CHW referrals, supporting caregiver reports of bypassing CHWs.

#### Step 2: Diagnosis and correct administration of RAS

CHWs in Serenje demonstrated stronger adherence to severe malaria case management protocols than those in Chama and Mwinilunga. Most patients (95.3%) were tested with an RDT, with positivity highest in Chama (89.0%) and Serenje (90.6%), and lowest in Mwinilunga (60.0%) (Fig 4) (S2 **Error! Reference source not found.**). Among those RDT positive, 104/106 (98.1%) were administered RAS in Serenje and 65/97 (67.0%) in Chama. In Mwinilunga, RAS was administered to 13 children who tested RDT-negative, resulting in a total of 49 administrations compared to 36 RDT-positive cases (136.1%). Among those given RAS, most were dosed correctly, except in Mwinilunga, where 15/49 (30.6%) were underdosed relative to age/weight guidelines [25].

**Fig 4.**
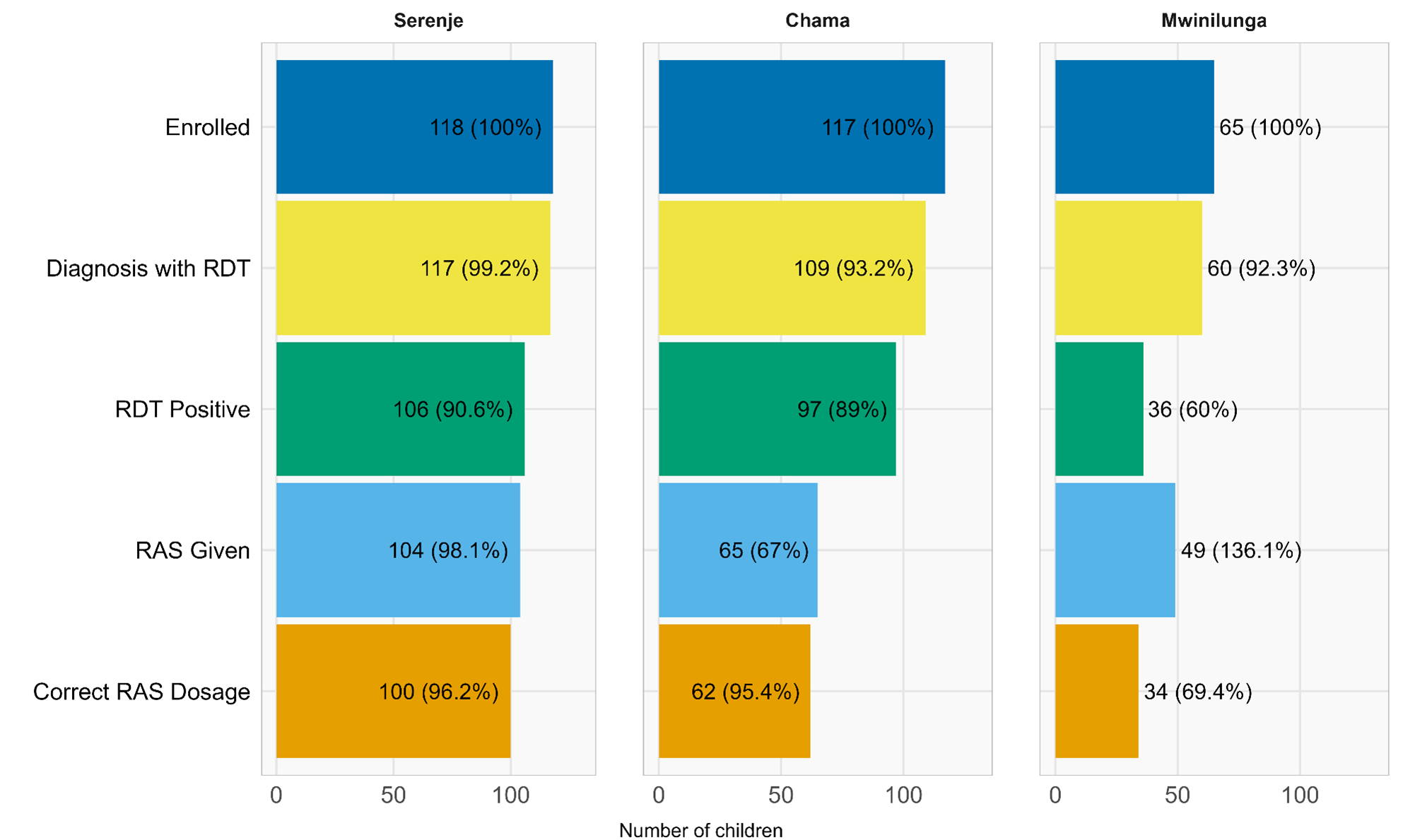
Step 2 - Diagnosis and RAS administration funnel chart by district. Funnel charts show the number and percentage of children progressing through each step of diagnosis and RAS administration in each district. Each stage’s denominator reflects the number of children from the preceding step, illustrating where drop-offs or deviations from the expected cascade occur. NOTE: RDT=rapid diagnostic test, RAS=rectal artesunate

Qualitative data highlighted CHW confidence in administering RAS and caregiver trust as key facilitators in Serenje and Chama. A CHW described a life-saving case that transformed community perceptions: *“It was a miracle, and we would have buried that child. People have come to believe RAS because of that child”. (CHW 1 IDI, Serenje).* Barriers included CHW unavailability and frequent stockouts of RAS. Some CHWs adapted by improvising PPE: *“If you don’t have gloves, you can use an empty plastic for sugar to put to your hand and administer RAS.” (CHW 2 IDI, Chama)*.

aSARA findings reinforced these patterns. From August to September of 2023, RAS availability was 75% in Serenje, 86% in Chama, but only 6.5% in Mwinilunga (S3 Table). Stockouts in Mwinilunga were prolonged, with 74.2% of CHWs reporting a RAS stockout in the past month, compared to 44.8% in Chama and 16.7% in Serenje. Referral slip and PPE shortages were also most severe in Mwinilunga.

### Step 3: Immediate referral and referral completion

Among children followed up at the health facility, 80.1% were recorded in facility logbooks, with Chama having the highest referral completion at 95.3%, followed by Mwinilunga at 75.8%, and finally Serenje at 71.1% (Fig 5). Same-day referral was highest in Mwinilunga (91.5%) despite the absence of an ETS (S4 Table). Serenje had the lowest same-day referral rate (80.2%), though transport options were more available.

**Fig 5.**
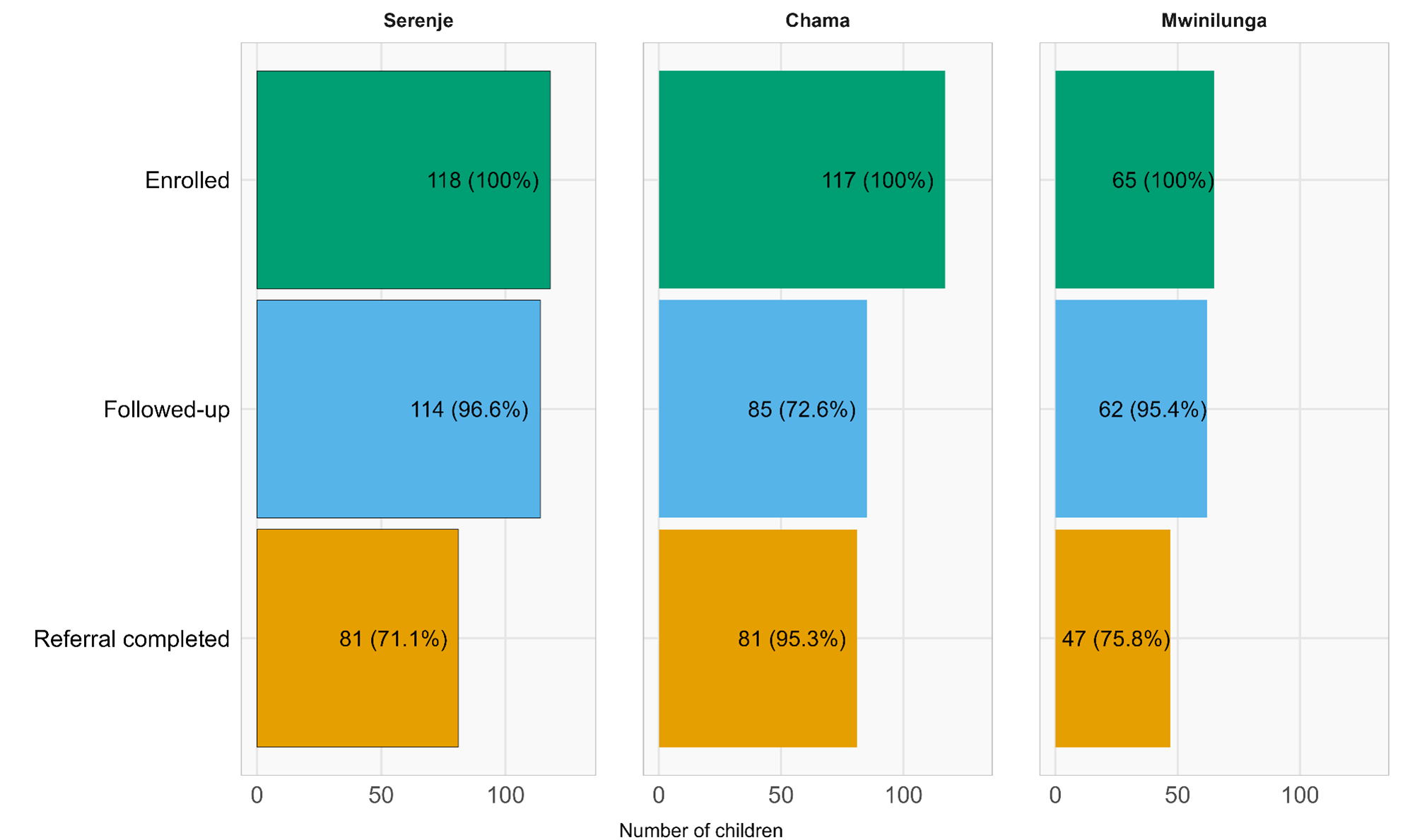
Step 3 - Immediate referral funnel chart by district. Funnel charts showing the proportion of enrolled children with suspected severe malaria who were followed up by the study team at the referral health facilities and completed referral in each district.

Qualitative findings showed that CHW insistence and caregiver trust drove referral completion. A caregiver described acting immediately when advised that RAS was only a temporary treatment: *“[The CHW] saw that the child was fitting… They put RAS on the child and said start off immediately; don’t go home, just go straight to Chibale clinic”. (Caregiver 2 IDI, Chama)*.

Where ETS was available, it was highly valued, and CHWs often helped to arrange alternative transport in its absence. In Mwinilunga, most families walked to the health facility, with some travelling 3–17 km. Transport shortages were compounded by poor roads, rugged terrain, and damaged bicycles as a traditional leader explained: *“We have no transport… You find a patient can even blackout and fall down”. (Traditional leader 1 IDI, Mwinilunga).* Household responsibilities and financial constraints also delayed or prevented referrals. Mothers, often the sole carers, sometimes postponed travel to secure food or care for other children. A health officer noted: *“They will even take time—sometimes even three days—they are still home”. (Health Officer 1 IDI, Chama)*.

Across districts, Serenje benefited from stronger community support systems, while Mwinilunga faced the greatest geographic and transport barriers. Chama’s experience fell between these two, with frequent transport challenges but stronger trust in referral advice than Mwinilunga.

### Step 4: Prompt treatment with injectable AS and complete treatment with AL

Most children with severe malaria who completed the trip to a referral facility received injectable AS, though follow-on AL provision was inconsistent—particularly in Mwinilunga, where over half (51.6%) of children were not given AL before discharge (Fig 6). Across the districts, 89.8% of severe malaria cases received both Inj AS and AL (S5 Table). Stockouts were rare for Inj AS but more common for AL during the study period.

**Fig 6.**
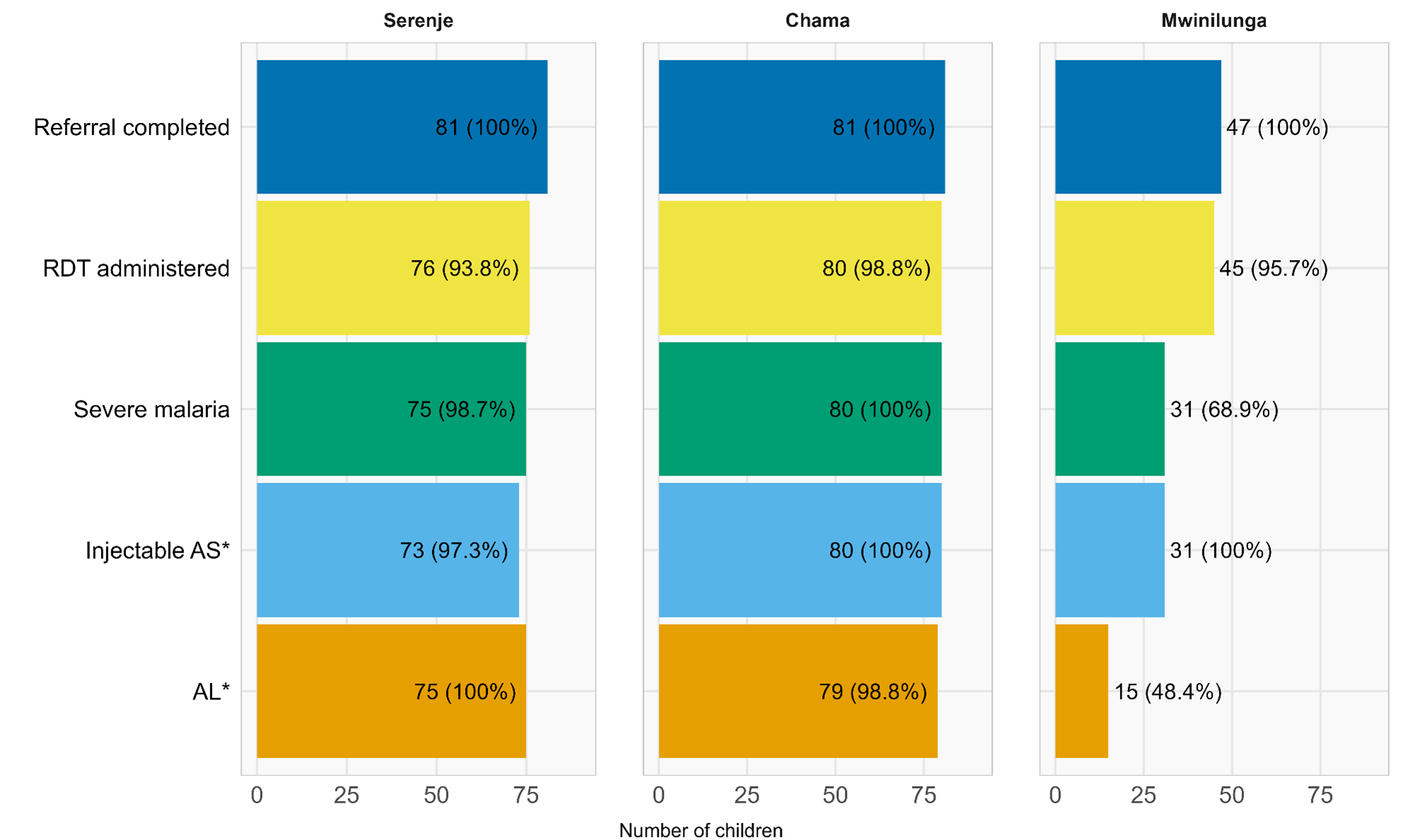
Step 4 - Complete treatment funnel chart by district. Funnel charts showing progression of children with suspected severe malaria who completed referral and received treatment in each district. Note: RDT = rapid diagnostic test, AS = artesunate, AL = artemether lumefantrine, * Proportions for injectable AS and AL are relative to those diagnosed with severe malaria at the referral health facility.

Qualitative interviews showed that referral slips from CHWs helped caregivers navigate facilities and receive timely Inj AS. A caregiver recounted her experience, *“We met the doctor and we gave him the referral letter… Then he said the child was very sick and needed urgent medical attention”. (Caregiver 2 IDI, Chama).* Most caregivers reported completing the AL course at home, driven by clear counseling or fear of malaria recurrence. A FGD participant explained: *“If you take half, you are going to kill half of the parasites… and they will multiply”. (FGD Participant 3, Mwinilunga).* However, some caregivers stopped treatment early once symptoms improved, saved doses for future use, or shared them with other sick children. Negative health worker attitudes also discouraged some caregivers from returning to complete treatment.

District-level differences mirrored earlier steps: Serenje showed stronger facility readiness and community adherence, Chama faced mixed commodity and staffing gaps, and Mwinilunga experienced the highest AL shortages despite strong self-reported completion at home.

## Discussion

This study assessed the implementation of Zambia’s RAS intervention package under routine conditions, examining real-world progression through the severe malaria cascade of care and the facilitators and barriers influencing each step. Quantitative findings revealed key points of disengagement after seeking care with a CHW, particularly during RAS administration, referral completion, and continuity of care, while qualitative insights revealed the systemic and behavioural factors shaping these outcomes.

Most caregivers interviewed were mothers, who typically serve as the primary decision-makers for child health. Caregivers and providers broadly viewed RAS as a valuable, fast-acting intervention that stabilized critically ill children and allowed time to reach referral facilities. Its community-level acceptability and perceived effectiveness were important facilitators, consistent with other settings [16–18, 26]. However, mothers’ treatment-seeking was constrained by complex social and structural factors. While responsible for recognizing danger signs and initiating care, they often had limited authority and financial autonomy, and had to balance household duties with the urgency of seeking care [9, 27–29]. In some districts, trust in CHWs and previous community engagement activities which had addressed some of the key social determinants that prevented timely treatment-seeking were reported as having supported earlier care seeking, but in some locations persistent gendered barriers continued to shape differences in timely care-seeking [20, 21].

The experience in Serenje, where RAS was piloted with intensive CHW and health worker training, robust supervision, community engagement activities and a strengthened supply chain, demonstrates the potential of a fully supported implementation [20, 21]. By contrast, Chama and Mwinilunga received less consistent support, contributing to some improper RAS use, including administration despite negative RDT results and under-dosing. Qualitative findings suggested that these deviations were often driven by CHWs’ perceptions of clinical urgency and concerns about delays to referral when children appeared critically ill. Similar patterns have been documented elsewhere, where limited training and supervision, stockouts, and perceived urgency can lead CHWs to deviate from guidelines under pressure [30–33]. In some cases, scarce supplies may prompt under-dosing as a rationing strategy [11].

While referral completion has often been identified as a critical bottleneck in severe malaria care, children from this study did not reach subsequent steps at multiple points along the cascade, with important variation across districts. In Serenje, losses accumulated steadily following RAS administration, including during referral and facility-based care. In Chama, the largest drop-off occurred earlier, at the point of RAS administration following a positive RDT. In Mwinilunga, substantial attrition occurred after referral, particularly at the stage of completing treatment with artemether–lumefantrine. These findings suggest that no single step universally represents the weakest link; rather, gaps in care reflect district-specific implementation challenges across both community and facility levels.

Referral completion remained a vulnerable step in the cascade of care, particularly from the perspective of caregivers. While immediate clinical improvement following RAS sometimes motivated families to complete referral, others interpreted recovery as a sign that further care was unnecessary, a finding consistent with results from the CARAMAL study [11, 12]. Practical constraints, including competing caregiver responsibilities, lack of food, and long travel distances further shaped referral decisions, as observed in other settings [34–36]. To mitigate these barriers, Zambia introduced a package of community-based supports, including ETS, food banks, and savings schemes. These interventions were highly valued but proved fragile as external support was reduced, leading many to lapse due to funding gaps and weak oversight. Without reliable community-managed financial, material, and transport supports to act quickly in a child’s illness, referral completion remains contingent on household resources and circumstances rather than health system design [37, 38].

Even when caregivers reached a health facility, full treatment was not guaranteed. Participants described inconsistent care, frequent stockouts, and occasional negative health worker attitudes—factors known to undermine trust and deter future care-seeking [33, 39, 40]. While injectable AS is now more widely available due to Zambia’s MAM@Scale initiative, challenges remain in administering follow-on treatment with AL [20, 21]. Despite no reported AL stockouts, qualitative findings suggested that gaps in follow-on treatment may reflect limited awareness of AL protocols, concerns about caregiver adherence, or inefficiencies in medicine distribution, as observed in other settings [33, 41, 42]. Variability in care quality likely reflects broader systemic issues such as staff burnout, weak supervision, and resource constraints [33, 39, 43]. Strengthening supportive supervision, mentorship, and clearly delineating CHW and facility staff responsibilities may help address these bottlenecks [44, 45].

Zambia’s phased rollout of RAS underscores the importance of contextualized implementation. Serenje illustrates best practices, while scale-up districts reveal gaps due to reduced support. Tailoring national policy with district-level investments in training, commodities, and supervision is essential. Ultimately, RAS effectiveness depends on the health system and community context in which it is delivered. Zambia’s efforts to address structural barriers through community savings schemes and food banks demonstrate both the promise and realities of these approaches. While community leadership was central to their design, sustaining these systems required intensive initial coaching, mentoring, and resource support from DHMTs and light-touch ongoing support. Where such support, either initial or ongoing, was not available, some community systems failed to become fully established or effectively community-led and managed and hence were unable to influence referral completion and outcomes. Locally led models, when coupled with reliable financing, integration into governance structures, and sustained DHMT engagement, are more likely to endure and enable caregivers—particularly mothers, who often recognize danger signs first—to act quickly with the support of household decision-makers when a child becomes severely ill. Without these investments, simply distributing RAS risks poor outcomes; in a small number of study facilities, RAS even expired because health staff were unaware of its intended use.

Despite these challenges, many caregivers showed strong motivation to complete referral—especially when they understood that RAS was only the first step. Many were willing to overcome substantial obstacles when equipped with accurate information and practical support. This highlights the importance of pairing awareness raising with sustainable community-based systems that empower families to complete the cascade of care.

This study has several limitations. First, the design captured only children who sought care from CHWs and therefore could not quantify the proportion of children with suspected severe malaria who never entered the cascade of care. Second, follow-up was incomplete; some families could not be located due to migration or the remoteness of villages, which limited the ability to ascertain referral completion and survival outcomes for all cases. Third, reliance on facility records introduced potential biases from incomplete or inaccurate documentation, and survival status was not consistently verified through community follow-up. Finally, qualitative findings may not be generalizable beyond the three study districts, although triangulation across respondent types and regions strengthens their credibility. These limitations highlight the need for caution in interpreting absolute estimates, while reinforcing the value of identifying system-level barriers and facilitators to RAS implementation.

As countries consider RAS introduction or scale-up, Zambia’s experience demonstrates that the intervention must be embedded within the broader continuum of severe malaria care rather than implemented in isolation. Newly released guidelines outline practical steps for doing so, including integrating RAS into national protocols, ensuring CHW competency through training and supervision, addressing the key social determinants that prevent timely treatment-seeking, maintaining reliable supply chains, and strengthening referral and transport systems [46]. Countries seeking to scale up RAS should consider adapting these recommendations to their own health system capacities and community contexts to maximize its life-saving potential.

## Conclusion

This study shows that the success of RAS as a life-saving intervention for severe malaria depends not only on the drug itself but also on the systems that support its delivery and use. While RAS can stabilize a child and buy time for referral, its impact is limited without strong community engagement, reliable supply chains, trained health workers, and accessible follow-up care. Zambia’s experience highlights the importance of pairing national policies with local investments in training, supervision, caregiver support and broader community engagement. For RAS to reach its full potential, it must be part of a broader effort to strengthen the entire cascade of care for severe malaria.

## List of abbreviations

ACT: rArtemisinin-based Combination Therapy
AL: rArtemether-Lumefantrine
AS: rArtesunate
aSARA: rAdapted Service Availability and Readiness Assessment
CHW: rCommunity Health Worker
CARAMAL: rCommunity Access to Rectal Artesunate for Malaria
DHMT: rDistrict Health Management Team
ETS: rEmergency Transport System
Inj AS: rInjectable Artesunate
IRB: rInstitutional Review Board
MAM@Scale: rMothers Against Malaria @ Scale
NMEC: rNational Malaria Elimination Centre
RAS: rRectal Artesunate
RDT: rRapid Diagnostic Test
WHO: rWorld Health Organization

## Declarations

## Acknowledgements

We are deeply grateful to the community health workers, health facility staff, and caregivers who participated in this study and generously shared their time and experiences. We thank the dedicated data collection teams in Zambia for their commitment under challenging conditions. We also acknowledge the support of our in-country partners, the Ministry of Health, PATH, and the National Malaria Elimination Centre, whose collaboration made this research possible.

## Funding

Bill & Melinda Gates Foundation through PATH MACEPA. The funder of the study had no role in study design, data collection, data analysis, data interpretation, or writing of the report. The corresponding author had full access to all the data in the study and had final responsibility for the decision to submit for publication.

## Ethics approval and consenting to participate

Ethical approval was obtained from the University of Zambia Biomedical Research Ethics Committee (REF. NO. 3941-2023), Tulane University Human Research Protection Program IRB (REF. NO. 2023-050), and PATH Office of Research Affairs IRB (REF.NO. RREV-001410). All participants were above 16 years of age (age of consent in Zambia) and provided written informed consent followed by verbal consent prior to their inclusion in the study.

## Data availability

The data underlying this study are derived from routine community health worker logbooks, health facility records, and qualitative interviews collected as part of an evaluation of RAS implementation conducted in collaboration with the Zambia Ministry of Health. These data contain sensitive health information and are owned and managed by the Zambia Ministry of Health (through the National Malaria Elimination Centre, NMEC) and implementing partners. As such, they cannot be shared publicly by the authors.

Qualified researchers may request access from the Zambia Ministry of Health at or via NMEC at. Requests will be evaluated in line with Ministry policies, applicable ethical approvals, and data-use agreements. The authors accessed the data under formal approvals and had no special access privileges; other researchers may request access through the same process.

## Notes

### Competing Interest Statement

The authors have declared no competing interest.

### Author Declarations

University of Zambia Biomedical Research Ethics Committee (REF. NO. 3941-2023), Tulane University Human Research Protection Program IRB (REF. NO. 2023-050), and PATH Office of Research Affairs IRB (REF.NO. RREV-001410). All provided ethical approval for this work.

