## Supplementary material for "Implementation Adherence and Operational Challenges of Rectal Artesunate for Severe Malaria in Zambia: A Mixed-Methods Study": S1

### S1 Appendix

S1 Table . Enrolment and follow-up status of severe malaria patients across the study districts

| Follow-up status | Serenje^1^ | Chama^1^ | Mwinilunga^1^ | Overall^1^ |
| --- | --- | --- | --- | --- |
| **Children with suspected severe malaria enrolled** | 118 | 117 | 65 | 300 |
| **Health facility follow-up attempted (% of enrolled)** | 114 (96.6%) | 85 (72.6%) | 62 (95.4%) | 261 (87.0%) |
| **Patient found in register (% of attempted)** | 81 (71.1%) | 81 (95.3%) | 47 (75.8%) | 209 (80.1%) |
| **Home follow-up attempted (% of enrolled)** | 100 (84.7%) | 51 (43.6%) | 51 (78.5%) | 202 (67.3%) |
| **Family found at home (% of attempted)** | 80 (80.0%) | 49 (96.1%) | 39 (76.5%) | 168 (83.2%) |
| **Complete follow-up (both health facility + home)** | 58 (49.2%) | 48 (41.0%) | 32 (49.2%) | 138 (46.0%) |
| ^1^n (%) | | | | |

S2 Table. Second step in the cascade of care for severe malaria: Diagnosis and correct administration of RAS

|  | Serenje |  | Chama |  | Mwinilunga |  | All |  |
| --- | --- | --- | --- | --- | --- | --- | --- | --- |
| Step 2. Diagnosis and correct administration of RAS | **n/N** | **%** | **n/N** | **%** | **n/N** | **%** | **n/N** | **%** |
| Diagnosis with RDT: Yes | 117/118 | 99.2 | 109/117 | 93.2 | 60/65 | 92.3 | 286/300 | 95.3 |
| RDT result: Positive | 106/117 | 90.6 | 97/109 | 89.0 | 36/60 | 60.0 | 239/286 | 83.6 |
| RAS given: Yes | 104/106 | 98.1 | 65/97 | 67.0 | 49/36 | 136.1 | 218/239 | 91.2 |
| Correct RAS dosage* | 100/104 | 96.2 | 62/65 | 95.4 | 34/49 | 69.4 | 196/218 | 89.9 |
| Referral slip given: Yes | 118/118 | 100.0 | 113/117 | 96.6 | 55/65 | 84.6 | 286/300 | 95.3 |

(200mg) for children over 3 years to less than 6 years old.

S3 Table. CHW aSARA results of commodity availability, recent stockouts, and stockout duration among CHWs across the study districts

| CHW Commodity Availability | Serenje  N = 24^1^ | Chama  N = 29^1^ | Mwinilunga  N = 31^1^ | Overall  N = 84^1^ |
| --- | --- | --- | --- | --- |
| **Availability** |  |  |  |  |
| RDT | 20 (83.3%) | 29 (100.0%) | 29 (93.5%) | 78 (92.9%) |
| AL | 19 (79.2%) | 28 (96.6%) | 22 (71.0%) | 69 (82.1%) |
| RAS | 18 (75.0%) | 25 (86.2%) | 2 (6.5%) | 45 (53.6%) |
| PPE | 21 (87.5%) | 18 (62.1%) | 3 (9.7%) | 42 (50.0%) |
| Referral slips | 20 (83.3%) | 29 (100.0%) | 6 (19.4%) | 55 (65.5%) |
| **Recent stockouts (in the past 4 weeks)** |  |  |  |  |
| RDT | 4 (16.7%) | 5 (17.2%) | 9 (29.0%) | 18 (21.4%) |
| AL | 4 (16.7%) | 3 (10.3%) | 18 (58.1%) | 25 (29.8%) |
| RAS | 4 (16.7%) | 13 (44.8%) | 23 (74.2%) | 40 (47.6%) |
| PPE | 5 (20.8%) | 15 (51.7%) | 25 (80.6%) | 45 (53.6%) |
| Referral slips | 3 (12.5%) | 6 (20.7%) | 25 (80.6%) | 34 (40.5%) |
| **Stockout duration** |  |  |  |  |
| RDT |  |  |  |  |
| Less than 7 days | 1 (25.0%) | 0 (00.0%) | 0 (00.0%) | 1 (5.6%) |
| 7 to 14 days | 0 (00.0%) | 2 (40.0%) | 0 (00.0%) | 2 (11.1%) |
| More than 14 days | 3 (75.0%) | 3 (60.0%) | 9 (100.0%) | 15 (83.3%) |
| AL |  |  |  |  |
| Less than 7 days | 1 (25.0%) | 0 (00.0%) | 1 (05.6%) | 2 (08.0%) |
| 7 to 14 days | 0 (00.0%) | 0 (00.0%) | 0 (00.0%) | 0 (00.0%) |
| More than 14 days | 3 (75.0%) | 3 (100.0%) | 17 (94.4%) | 23 (92.0%) |
| RAS |  |  |  |  |
| Less than 7 days | 1 (25.0%) | 3 (23.1%) | 1 (04.3%) | 5 (12.5%) |
| 7 to 14 days | 0 (00.0%) | 1 (07.7%) | 0 (00.0%) | 1 (02.5%) |
| More than 14 days | 3 (75.0%) | 9 (69.2%) | 22 (95.7%) | 34 (85.0%) |
| PPE |  |  |  |  |
| Less than 7 days | 0 (00.0%) | 1 (06.7%) | 0 (00.0%) | 1 (02.2%) |
| 7 to 14 days | 0 (00.0%) | 2 (13.3%) | 0 (00.0%) | 2 (04.4%) |
| More than 14 days | 5 (100.0%) | 12 (80.0%) | 25 (100.0%) | 42 (93.3%) |
| Referral slips |  |  |  |  |
| Less than 7 days | 0 (0.0%) | 6 (100.0%) | 1 (4.0%) | 7 (20.6%) |
| 7 to 14 days | 0 (00.0%) | 0 (00.0%) | 0 (00.0%) | 0 (00.0%) |
| More than 14 days | 3 (100.0%) | 0 (0.0%) | 24 (96.0%) | 27 (79.4%) |
| ^1^N (%) | | | | |

S4 Table. Third step in the cascade of care for severe malaria: Immediate referral and referral completion

|  | Serenje |  |  | | Chama | |  | Mwinilunga | |  | | Total | |
| --- | --- | --- | --- | --- | --- | --- | --- | --- | --- | --- | --- | --- | --- |
| Step 3. Immediate referral and referral completion | **n/N** | **%** |  | **n/N** | | **%** | **n/N** | | **%** | | **n/N** | | **%** |
| Health Facility follow-up |  |  |  |  | |  |  | |  | |  | |  |
| Not followed up | 4/118 | 3.4 |  | 32/117 | | 27.4 | 3/65 | | 4.6 | | 39/300 | | 13.0 |
| Attempted | 114/118 | 96.6 |  | 85/117 | | 72.6 | 62/65 | | 95.4 | | 261/300 | | 87.0 |
| Patient found in register |  |  |  |  | |  |  | |  | |  | |  |
| Found | 81/114 | 71.1 |  | 81/85 | | 95.3 | 47/62 | | 75.8 | | 209/261 | | 80.1 |
| Referral delay |  |  |  |  | |  |  | |  | |  | |  |
| Same day | 65/81 | 80.2 |  | 71/81 | | 87.7 | 43/47 | | 91.5 | | 179/209 | | 85.6 |
| Next day | 3/81 | 3.7 |  | 1/81 | | 1.2 | 0/47 | | 0.0 | | 4/209 | | 1.9 |
| Later than next day | 8/81 | 9.9 |  | 6/81 | | 7.4 | 2/47 | | 4.3 | | 16/209 | | 7.7 |
| Unknown | 5/81 | 6.2 |  | 3/81 | | 3.7 | 2/47 | | 4.3 | | 10/209 | | 4.8 |
| Caregiver indicated: Travel method |  |  |  |  | |  |  | |  | |  | |  |
| Emergency transport system | 16/78 | 20.5 |  | 17/49 | | 34.7 | 0/38 | | 0.0 | | 33/165 | | 20 |
| Car | 9/78 | 11.5 |  | 0/49 | | 0.0 | 1/38 | | 2.6 | | 10/165 | | 6.1 |
| Motorbike | 1/78 | 1.3 |  | 1/49 | | 2.0 | 1/38 | | 2.6 | | 3/165 | | 1.8 |
| Bus/public transportation | 1/78 | 1.3 |  | 0/49 | | 0.0 | 1/38 | | 2.6 | | 2/165 | | 1.2 |
| Other bicycle | 28/78 | 35.9 |  | 25/49 | | 51.0 | 1/38 | | 2.6 | | 54/165 | | 32.7 |
| Walk | 23/78 | 29.5 |  | 6/49 | | 12.2 | 34/38 | | 89.5 | | 63/165 | | 38.2 |
| Travel time to health facility (median and SD) | 60.0 | 82.5 |  | 60.0 | | 45.0 | 90.0 | | 81.3 | | 60.0 | | 75.0 |

IQR = interquartile range, Other bicycle includes personal bicycle, borrowed bicycle, or hired bicycle

S5 Table. Fourth step in the cascade of care for severe malaria: Prompt and complete treatment with injectable artesunate and AL

|  | Serenje |  | Chama |  | Mwinilunga |  | All |  |
| --- | --- | --- | --- | --- | --- | --- | --- | --- |
| Step 4. Prompt and complete treatment with injectable artesunate and AL | **n/N** | **%** | **n/N** | **%** | **n/N** | **%** | **n/N** | **%** |
| Health facility diagnosis: Malaria | 76/81 | 93.8 | 80/81 | 98.8 | 45/47 | 95.7 | 201/209 | 96.2 |
| Level of malaria: Severe | 75/76 | 96.2 | 80/80 | 100.0 | 31/45 | 68.9 | 186/201 | 92.5 |
| Treatment given: Yes | 76/81 | 98.7 | 80/81 | 98.8 | 45/47 | 95.7 | 201/209 | 96.2 |
| Health facility treatment: |  |  |  |  |  |  |  |  |
| Injectable AS | 73/75 | 97.3 | 80/80 | 100.0 | 31/31 | 100.0 | 184/186 | 98.9 |
| AL | 75/75 | 100.0 | 79/80 | 98.8 | 15/31 | 48.4 | 169/186 | 90.9 |
| Both | 73/75 | 97.3 | 79/80 | 98.8 | 15/31 | 48.4 | 167/186 | 89.8 |
| At home follow-up: Attempted | 100/118 | 84.7 | 51/117 | 43.6 | 51/65 | 78.5 | 202/300 | 67.3 |
| At home follow-up: Family found | 80/100 | 80.0 | 49/51 | 96.1 | 39/51 | 76.5 | 168/202 | 83.2 |
| Caregiver indicated: |  |  |  |  |  |  |  |  |
| Received AL: Yes | 76/80 | 95.0 | 48/49 | 98.0 | 39/39 | 100.0 | 163/168 | 97.0 |
| Completed AL: Yes | 76/76 | 100.0 | 48/48 | 100.0 | 38/39 | 97.4 | 162/163 | 99.4 |
| CHW follow-up: Yes | 80/80 | 100.0 | 40/49 | 81.6 | 33/39 | 84.6 | 153/168 | 91.1 |

Note: AS = artesunate, AL = artemether lumefantrine, CHW = community health worker

S6 Table. Number of interviews by respondent type across the study districts or at the national level

| Respondent Type | Serenje | Chama | Mwinilunga | National | Total |
| --- | --- | --- | --- | --- | --- |
| In-depth Interviews (IDIs) |  |  |  |  |  |
| Caregiver | 15 | 11 | 5 | 0 | 31 |
| Community health workers (CHWs) | 15 | 15 | 7 | 0 | 37 |
| District/Provincial Officers | 1 | 3 | 3 | 0 | 7 |
| Emergency transport providers (ETP) | 2 | 4 | 0 | 0 | 6 |
| Health facility staff | 3 | 2 | 2 | 0 | 7 |
| National staff | 0 | 0 | 0 | 3 | 3 |
| Traditional leader | 3 | 2 | 2 | 0 | 7 |
| Total IDIs | 39 | 37 | 19 | 3 | 98 |
| Focus Group Discussions (FGDs) |  |  |  |  |  |
| Focus group discussion | 4 | 2 | 4 | 0 | 10 |

S7 Table. Summary table of the main themes discovered regarding caregiver treatment seeking and the cascade of care for severe malaria

|  | Main Themes Discovered | |
| --- | --- | --- |
| Critical steps in the cascade of care | **Facilitators** | **Barriers** |
| Step 1. Caregiver treatment-seeking | - Trust in CHW capabilities and service | - Caregiver decision-making and delays in treatment-seeking |
| Step 2. Diagnosis and administration of RAS | - CHW belief in self-efficacy to administer RAS - Perceived effectiveness of RAS | - Limited CHW availability - Stockouts of malaria commodities |
| Step 3. Immediate referral | - Urgency and trust in referral advice - Health systems support (i.e., transport or financial assistance) | - Transportation and geographic constraints - Caregiver responsibilities and financial limitations |
| Step 4. Complete treatment | - Prioritization and continuity of care for referred patients - Fear that malaria may come back | - Lack of essential resources - Negative health worker attitudes - Caregiver perception and non-adherence to AL treatment |
